# Virtual peer role-play during COVID-19 pandemic for teaching medical students how to break bad news

**DOI:** 10.1101/2021.03.25.21254288

**Authors:** Jebrane Bouaoud, Pierre Saintigny

## Abstract

In order to cope with the SARS-CoV-2 pandemic and meet with the educational needs of medical students, we have evaluated the virtual peer role-plays (VPRP), an innovative approach to teach breaking bad news communication skills to medical students. Three scenarios of relational simulation were successively proposed to 237 medical students divided in 10 groups, each supervised by two teachers. Pre- and post-VPRP questionnaires were submitted to evaluate students’ satisfaction. The response rate of the pre- and post-VPRP questionnaires were 89% and 52% respectively. Two-thirds of the students had never participated in a peer role-play session. Most students had low level of confidence in breaking bad news communication and were motivated to participate to the VPRP session. Students’ satisfaction on VPRP session regarding quality (realism, organization), interest, perceived benefits was very positive. In conclusion, VPRP are feasible, of low cost (no material is required), applicable to other healthcare students and is relevant to the growth of teleconsultation.

## INTRODUCTION

Breaking bad news communication is one of the most challenging skills to teach to medical students [1]. Among simulation-based training methods, peer role-play is usually appreciated and seems improve communication skills in medical students [2].

SARS-CoV-2 pandemic has led to public health measures to prevent the virus spreading that significantly impacted medical students’ curriculum including formal lectures, directed studies, and practical work [3]. In particular, the directed studies to teach communication skills on how to break bad news are not anymore possible as they require simulation-based face-to-face peer-role play in small teaching rooms including 20-25 students. Because breaking bad news to patients is an important responsibility of physicians and a source of anxiety among medical students, and to meet students’ educational needs, we decided to maintain a training session for breaking bad news communication in the oncology field by proposing innovative virtual peer role-plays (VPRP).

## MATERIALS AND METHODS

Each VPRP session were supervised by two teachers (medical practitioners in oncology) and included 20-25 undergraduate medical students and lasted two hours. After reviewing the learning objectives, teachers reminded general principles of communication and relational skills such as learning to listen, working on one’s own emotions and building a quality relationship with the patient. Then a 30 minutes brainstorming was done around a challenging clinical situation (newly diagnosed metastatic pancreatic adenocarcinoma in a 65-year-old patient). The following question “how to give hope to a patient in a seemingly hopeless situation” was discussed, allowing to provide tips and advices to the students. Finally, the actual VPRP began. Three relational simulation scenarios were proposed: the first two were teleconsultations and the third one was a phone call (Supplemental Material 1-3). On a voluntary basis, two students participated to the first VPRP session. In two virtual rooms, isolated from the rest of the group, one of the teachers presented the scenario to the student playing the patient’s role, while the other teacher presented the scenario to the second student playing the doctor’s role. After 5 minutes of briefing, both students performed the VPRP in front of all the other students and teachers. Only the two “actor” students had their webcams and microphones on. The students were reminded that they could “Stop” the play in case they felt uncomfortable and/or to get some advices and to “Rewind” the peer role play to try another approach. After the VPRP session, the debriefing was conducted by the two teachers, starting with the student who played the “doctor,” and then the student who played the “patient”, then the other students. Several points were discussed such as their feelings during the play, and tips were given to feel more comfortable with breaking bad news communication while preserving the attention and the confidence of the patient. The second and third VPRP session were then successively played by two other couple of students playing the roles of the “patient or trusted support person” or the “doctor”. The two teachers wrapped-up the session by summarizing key points discussed during the three debriefings.

Before the VPRP session, each students was asked to complete a 5 items self-questionnaire to collect basic students characteristics (age, gender), as well as the following information: i-whether they had any previous experience in a peer role-play to develop communication skills for breaking bad news, ii-whether they felt confident regarding their ability to communicate breaking bad news to a patient (six-point likert scale) and iii-whether they were motivated to participate in the VPRP session (six-point likert scale).

Based on the level 1 of the Kirkpatrick training evaluation model (*i*.*e*., “Reaction”)[4], a post-VPRP session self-questionnaire (7-item questionnaire) was proposed to students.

## RESULTS

Overall, 10 VPRP sessions were organized simultaneously. A total of 237 students received the training. The response rate to the pre-VPRP session questionnaire was 89% (212/237 students). The mean age of the students was 22 years old (range 19-36) and the male/female sex ratio was 0.72. Two-thirds of the students had never participated to any peer role-play session. Most students had a low level of confidence (level 1-3, 70%; median=3) in breaking bad news but were motivated to participate to the VPRP session (level 4-6, 58%; median=5) (Supplemental Material 4).

The response rate to the post-VPRP session questionnaire was 52% (111/212 students). Most students (95%) rated the teaching as good to excellent and the scenario as very realistic (77%) (Supplemental Material 5). Most of them (67%) reported a great interest in VPRP session participation. Indeed, 86% of students stated that VPRP session was a helpful training in breaking bad news communication (Figure 1). In particular, most of them (80%) felt better prepared for breaking bad news communication after the VPRP session and recommended (89%) that the VPRP training session being maintained in the curriculum of future students (Supplemental Material 6).

**Figure 1.**
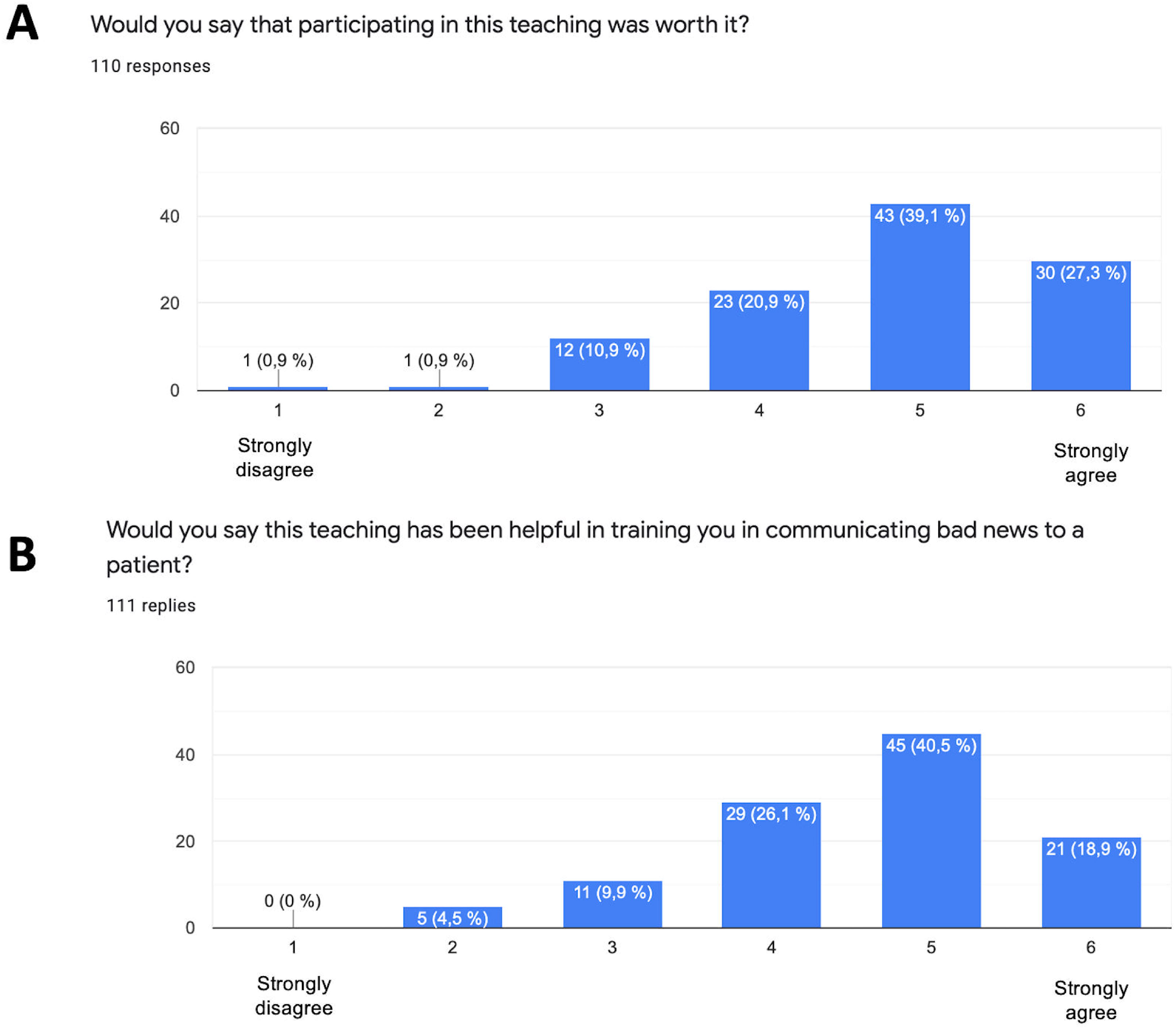
Histograms showing responses to the post-Virtual peer role-plays training session questionary, in particular regarding interest (**A**) and perceived benefits (**B**) of the training sessions.

## DISCUSSION

The value in medical curricula as well as major advantages in terms of cost-effectiveness of Peer role play have already been reported [5]. Interestingly, we found that students’ satisfaction on Virtual-PRP sessions regarding quality (realism, organization), interest, and perceived benefits was high. The VPRP session for breaking bad news communication offers several advantages. VPRP are low-cost (no material is required), and easily applicable to other healthcare students, which is of great interest. It allowed teaching continuation despite SARS-CoV-2-related public health measures and associated restrictions. Moreover, VPRP sessions address the specific challenge of teleconsultations and telephonic conversations. Our scenarios (two teleconsultations and one phone call) were particularly realistic given the actual pandemic. One fundamental aspect is the importance of structured pre-defined scenario as well as post-VPRP briefing to homogenize the learning skills. Involving physicians in the VPRP-sessions allowed us to ensure students supervision, support and feedbacks which are of great importance [6].

The main limitation of our VPRP sessions is that a minority of students actively played the role of “patient or trusted support person” or of the “doctor” (only 20% of students were “actors” during our sessions). This could be addressed by increasing the number of virtual rooms in each session. Overall, while the 52% rate of response to the post-VPRP questionnaire may be associated with a potential bias, we feel VPRP sessions are realistic, valuable and an effective method for learning breaking bad news communication skills. Future studies should evaluate the “learning” benefits of VPRP *i*.*e*., knowledge, skills, attitudes and perceptions (confidence, commitment).

## Data Availability

The data used and/or analyzed during the current study are available from the corresponding author on reasonable request.

## ACKNOWLEDGMENTS

The authors would thank Anaïs Saintigny (Pharmacy Student), Nathalie Borel (Medical Assistant), Professor Rode Gilles (Dean of the Université Claude Bernard Lyon 1, France) and Professor Debra Roter (Johns Hopkins Bloomberg School of Public Health) for their supports

## SUPPLEMENTARY MATERIAL

**Supplementary material 1**. Virtual peer role play session, scenario 1

**Supplementary material 2**. Virtual peer role play session, scenario 2

**Supplementary material 3**. Virtual peer role play session, scenario 3

**Supplementary material 4**. Responses to the pre-VPRP training session questionary

**Supplementary material 5**. Histograms showing responses to the post-Virtual peer role-plays training session questionary, in particular regarding the quality (**A**) and realism (**B**) of the training sessions.

**Supplementary material 6**. Histograms showing responses to the post-Virtual peer role-plays training session questionary, in particular regarding perceived benefits (**A**) and interest (**B**) of the training sessions.

