## Supplementary Material 1 for "Virtual peer role-play during COVID-19 pandemic for teaching medical students how to break bad news"

It is April 2020, in the midst of a health crisis related to the SARS-CoV-2 pandemic and the government has imposed a lockdown. Trips to access medical care are allowed with the condition of obtaining a certificate.

**Synopsis 1: Teleconsultation with concerning symptomatology… in the midst of a health crisis**

**You are a general practitioner**: you are seeing a 70-year-old patient for a teleconsultation to make an assessment of his spinal pain, not resulting from any trauma. The patient has a past history of lumbar osteoarthritis. You have already seen him for a teleconsultation 15 days ago. He had complained about pain, mostly in his back, not relieved by the usual antalgic treatment. He had lost 3 kg. The standard X-ray of the spine was considered normal. You had prescribed a level II antalgic treatment and scheduled a new appointment to reevaluate the patient’s condition. There is a possibility that an MRI is needed to understand the origin of his spinal pain, particularly looking for a tumoral process.

**You are the patient:** 70-year-old man, you are retired and live with your wife. You have suffered from lombar pain for a long time but lately, for the last 3 months, the pain has been higher in the spine and more intense than usual. At first, it progressed insidiously but is now practically permanent. This pain wakes you up at night. You have less appetite than usual, and you are losing weight. The pain medication prescribed by your general practician are only partially efficient. You are also very worried about the health crisis. One of your neighbors has contracted the Covid-19 virus and is currently being hospitalized in the Intensive Care Unit.
