## Supplementary Material 2 for "Virtual peer role-play during COVID-19 pandemic for teaching medical students how to break bad news"

It is April 2020, in the midst of a health crisis related to the SARS-CoV-2 pandemic and the government has imposed a lockdown. Trips to access medical care are allowed with the condition of obtaining a certificate.

**Synopsis 2: A concerning… or reassuring… vesicular image in a teleconsultation**

**You are a general practitioner:** a 56-year-old patient has an appointment with you for a teleconsultation regarding the results of his 6-month check-up which includes a hepatic ultrasound and the measurement of ACE (a serum marker) levels. The patient had surgery 6 years ago for a right hemicolectomy for a pT2 N1 colic adenocarcinoma. The ACE levels are normal, and the abdominal ultrasound report shows the presence of a non-complex biliary lithiasis.

**You are the patient:** 56-year-old man, you are in the company of your wife. You were diagnosed with colon cancer at your first screen test at the age of 50. After having gone through surgery, your doctor prescribed a check-up every 6 months. You are stressed because the latest ultrasound shows an abnormality in your gallbladder and you have read that gallbladder cancer have a bad prognosis and that they are hereditary.
