## Supplementary Material 3 for "Virtual peer role-play during COVID-19 pandemic for teaching medical students how to break bad news"

It is April 2020, in the midst of a health crisis related to the SARS-CoV-2 pandemic and the government has imposed a lockdown. Trips to access medical care are allowed with the condition of obtaining a certificate.

**Synopsis 3: an uncomfortable situation…in the middle of the night, in the midst of a pandemic, on the phone**.

**You are the on-call fellow:** after a hectic night in the emergency room, you can finally lay down to rest… but the phone suddenly rings in the on-call room and wakes you up at 4:30 AM… a nurse from the department of Rheumatology calls you to see if you could take care of a patient that you don’t know, who is in respiratory distress. When you walk into the patient’s room, you notice that the patient is unconscious, but some slow respiratory movements (gasps) are visible. You conclude the patient is in a state of shock, with a systolic blood pressure of 5, a bradycardia of 35 bpm and an oxygen saturation level of 85% with a high concentration oxygen mask (15 l/min). You quickly go through the patient’s medical chart: Mrs B. is a 68-year-old widow. According to her living will, a Do Not Resuscitate (DNR) order is in place. Indeed, she was diagnosed 3 months ago with metastatic pancreatic cancer that has spread to the liver for which she receives palliative care to reduce her symptoms. A hospitalization in her daughter’s home was also decided. The patient was admitted a few days ago with fever and she was diagnosed with COVID-19 on the basis of a Polymerase Chain Reaction (PCR) test and an abnormal image on the Magnetic Resonance Imaging (MRI). You need to call her emergency contact, her daughter, to let her know about the decline of her mother’s condition. Also, let her know that there is a risk of imminent death and you will start administering medication to make her comfortable.

**You are the patient’s daughter and her proxy:** 43 years old, single, you are the only child of Mrs B. and her proxy. These last few months have been hard. Your mother, who had not had any major health issues until now, was diagnosed with pancreatic cancer with hepatic metastases. It was a difficult diagnosis as many exams were required. During the last 3 months, she has lost 15 kg. You are having trouble accepting that the only recommended treatment is palliative care even though the doctors have explained that they think your mother cannot receive any chemotherapy because she is a frail elderly. You still have hope since her condition has gotten better after the drainage of her biliary tract. 3 days ago, you had insisted that she’d be brought to the ER by the fire department because of a fever, a cough and the loss of the sense of taste which reminded her of COVID-19 symptoms. It is 5:00 AM and the phone rings while you are sleeping. You pick up the phone and it is the hospital operator. The on-call fellow is transferred in…
