## Supplementary Material 4 for "Virtual peer role-play during COVID-19 pandemic for teaching medical students how to break bad news"

How would you rate your level of confidence in your ability to deliver bad news to a patient?

212 replies

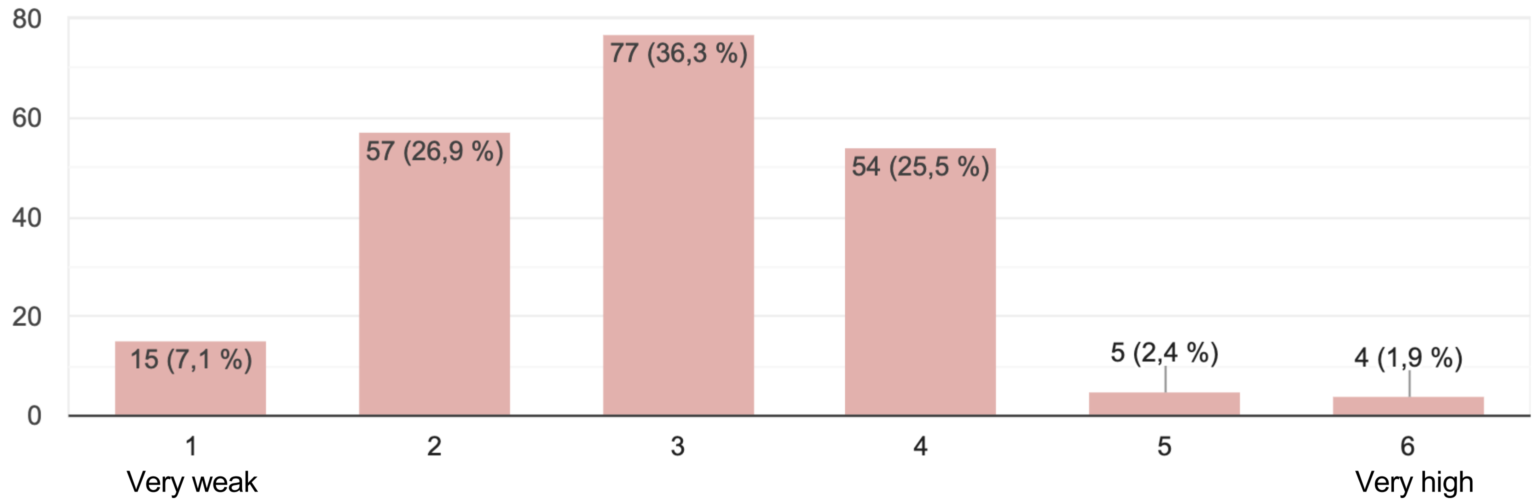


How would you judge your motivation to participate in this teaching?

211 replies

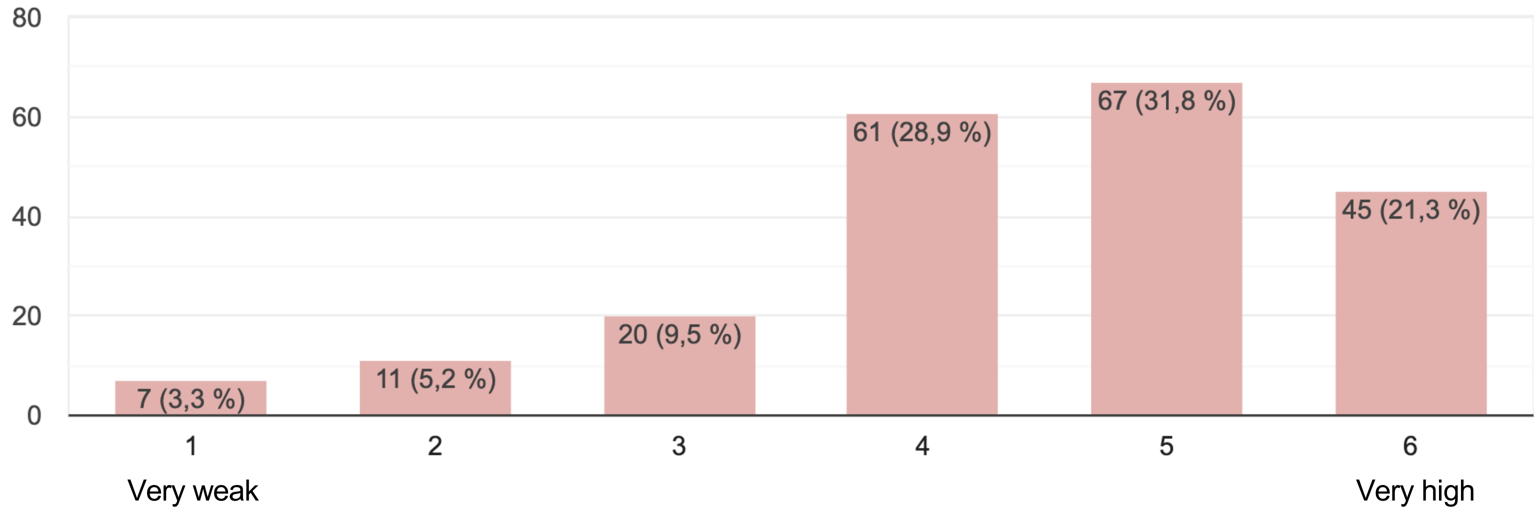
