## Supplementary Material 6 for "Virtual peer role-play during COVID-19 pandemic for teaching medical students how to break bad news"

**A**

Would you say that you feel like you are better prepared to deliver bad news to a patient?

110 responses

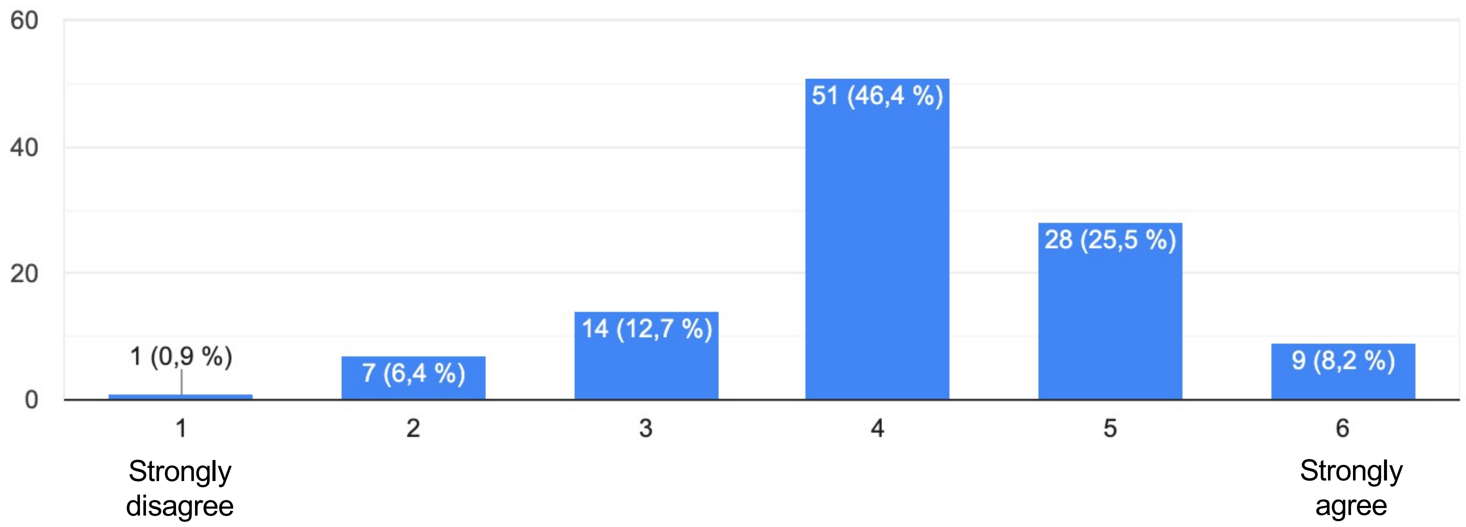


**B**

Would you recommend continuing this education next year?

111 replies

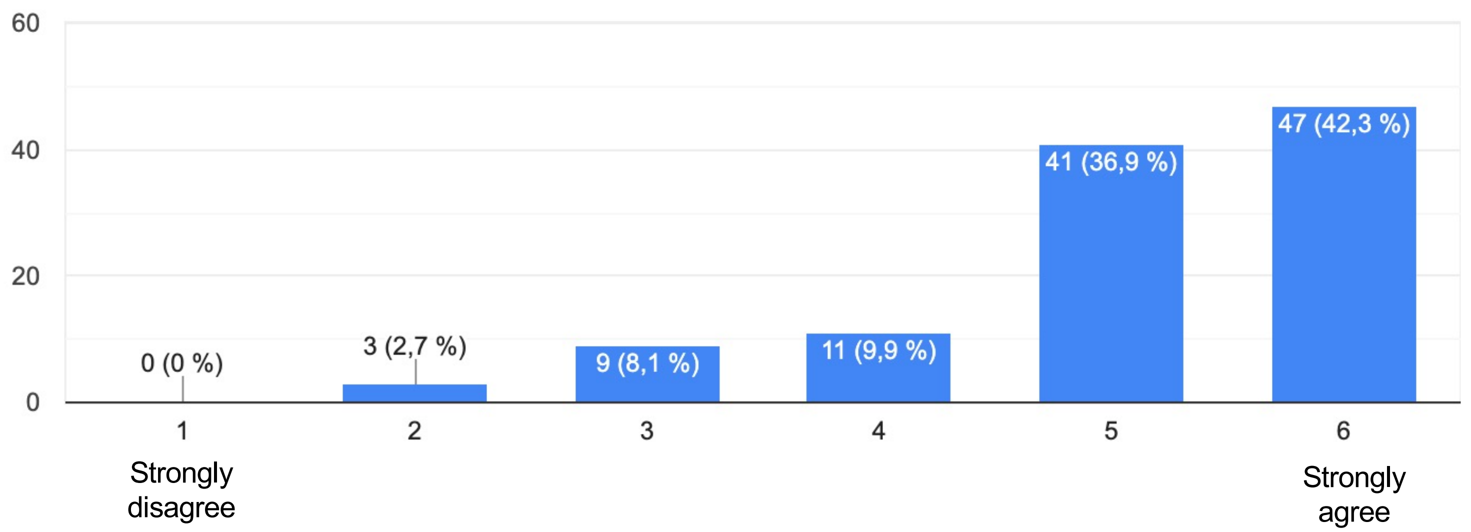
