## Supplementary Material 5 for "Virtual peer role-play during COVID-19 pandemic for teaching medical students how to break bad news"

**A**

Would you say that participating in this teaching was worth it?

110 responses

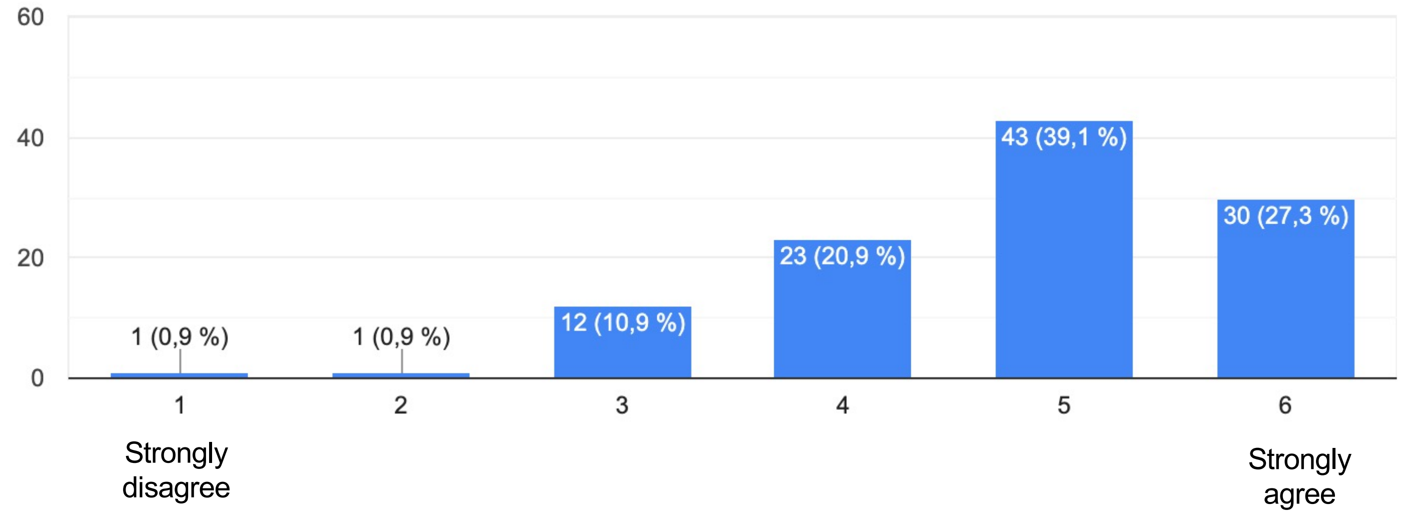


**B**

Would you say this teaching has been helpful in training you in communicating bad news to a patient?

111 replies

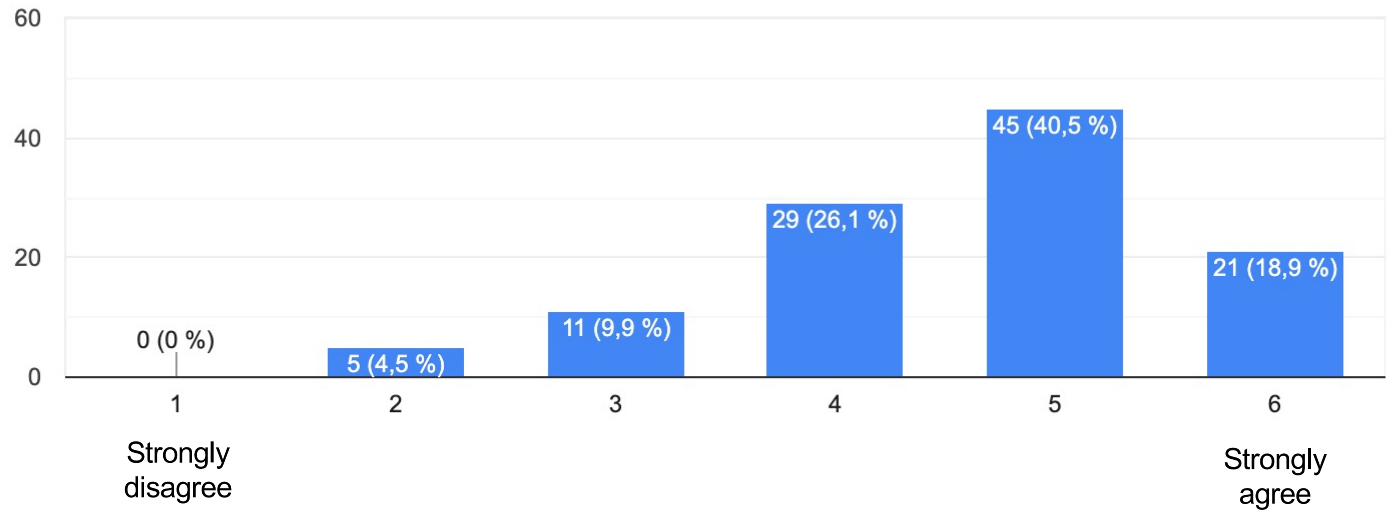
